# Epidemiological characteristics of COVID-19 cases in Italy and estimates of the reproductive numbers one month into the epidemic

**DOI:** 10.1101/2020.04.08.20056861

**Authors:** Flavia Riccardo, Marco Ajelli, Xanthi D Andrianou, Antonino Bella, Martina Del Manso, Massimo Fabiani, Stefania Bellino, Stefano Boros, Alberto Mateo Urdiales, Valentina Marziano, Maria Cristina Rota, Antonietta Filia, Fortunato Paolo D’Ancona, Andrea Siddu, Ornella Punzo, Filippo Trentini, Giorgio Guzzetta, Piero Poletti, Paola Stefanelli, Maria Rita Castrucci, Alessandra Ciervo, Corrado Di Benedetto, Marco Tallon, Andrea Piccioli, Silvio Brusaferro, Giovanni Rezza, Stefano Merler, Patrizio Pezzotti, for the COVID-19 working group

## Abstract

**Background:** In February 2020, a locally-acquired COVID-19 case was detected in Lombardia, Italy. This was the first signal of ongoing transmission of SARS-CoV-2 in the country. The outbreak rapidly escalated to a national level epidemic, amid the WHO declaration of a pandemic.

**Methods:** We analysed data from the national case-based integrated surveillance system of all RT-PCR confirmed COVID-19 infections as of March 24^th^ 2020, collected from all Italian regions and autonomous provinces. Here we provide a descriptive epidemiological summary on the first 62,843 COVID-19 cases in Italy as well as estimates of the basic and net reproductive numbers by region.

**Findings:** Of the 62,843 cases of COVID-19 analysed, 71.6% were reported from three Regions (Lombardia, Veneto and Emilia-Romagna). All cases reported after February 20^th^ were locally acquired. Estimates of R0 varied between 2.5 (95%CI: 2.18-2.83) in Toscana and 3 (95%CI: 2.68-3.33) in Lazio, with epidemic doubling time of 3.2 days (95%CI: 2.3-5.2) and 2.9 days (95%CI: 2.2-4.3), respectively. The net reproduction number showed a decreasing trend starting around February 20-25, 2020 in Northern regions. Notably, 5,760 cases were reported among health care workers. Of the 5,541 reported COVID-19 associated deaths, 49% occurred in people aged 80 years or above with an overall crude CFR of 8.8%. Male sex and age were independent risk factors for COVID-19 death.

**Interpretation:** The COVID-19 infection in Italy emerged with a clustering onset similar to the one described in Wuhan, China and likewise showed worse outcomes in older males with comorbidities. Initial R0 at 2.96 in Lombardia, explains the high case-load and rapid geographical spread observed. Overall Rt in Italian regions is currently decreasing albeit with large diversities across the country, supporting the importance of combined non-pharmacological control measures.

**Funding:** routine institutional funding was used to perform this work.

## INTRODUCTION

SARS-CoV-2 infection in humans causing clusters of severe pneumonia (1–3) was first detected in the city of Wuhan, China, in December 2019. This pathogen was confirmed as a new virus by the Coronavirus Study Group (CSG) of the International Committee on Taxonomy of Viruses. Based on phylogeny, taxonomy and established practice, they designated it as severe acute respiratory syndrome coronavirus 2 (SARS-CoV-2).(4) Although of probable zoonotic origin, human-to-human transmission is rapidly fuelling the spread of SARS-CoV-2 infections globally, with the main known routes of transmission being droplet and fomites.

The infection spread within China and rapidly across countries, with cases in Europe initially limited to small clusters in Germany, France (5,6) and the UK (7). On the 20^th^ of February 2020, the first case of locally acquired SARS-CoV-2 infection was diagnosed in Northern Italy in a critically ill, hospitalized young man with no travel history to known areas of viral circulation or link to a probable or confirmed COVID-19 case. Prior to this date, only three cases of COVID-19 had been reported in Central Italy, all with a travel history to Wuhan.

Following this unexpected finding, extensive contact tracing and testing of close contacts unveiled ongoing transmission in several municipalities of the Lombardia region (8,9). In subsequent days and weeks, case counts, and death tolls increased rapidly, at first in northern Italy and then in the rest of the country. The Italian government imposed increasingly strict physical distancing measures starting with the closure of 10 municipalities in the Lodi Province (Lombardia) and one in the Padua province (Veneto) on the 23^rd^ of February 2020. This culminated with a national lock-down on the 11th of March 2020. (10,11)

In this paper, we summarize key epidemiological findings from data on the first 62,843 confirmed COVID-19 cases in Italy, including 5,541 associated deaths, and initial findings on SARS-CoV-2 transmissibility across different regions.

## METHODS

### The Italian integrated COVID-19 surveillance system

With the aim to increase the understanding of the disease dynamics in Italy and support the planning of public health actions, a case-based surveillance system was established on the 27^th^ of February 2020, building on a previously existing surveillance system focused only on suspected and confirmed COVID-19 severe respiratory infections (12). The system contains data on all laboratory confirmed cases of COVID-19 as per the case definition published and regularly updated online by the European Centre for Disease Prevention and Control (ECDC) (13).

Laboratory confirmation by RT-PCR on nasopharyngeal swabs is performed at regional level, as previously described (9,14). From the beginning of the outbreak until the 1^st^ of March 2020, all initially confirmed cases were sent for final confirmation at the National Reference Laboratory in the Istituto Superiore di Sanità (ISS) and re-confirmed according to the World Health Organization guidelines using RT-PCR protocols described by Corman et al. (15) and by the US-CDC (16). Due to the high concordance (99%) among confirmation results with the engaged laboratories, the policy was then changed allowing selected Regions with demonstrated confirmation capacity to directly confirm COVID-19 cases (17). Data is collected daily using a secure online platform or, alternatively, received as individual datasets from the 19 regions and two Autonomous Provinces (AP) of the Italian territory, according to an increasingly harmonized track-record. Data collected includes information on: demographics, clinical severity, comorbidities, date of symptom onset, date of diagnosis, outcome, region of diagnosis and province of residence.

Clinical severity was defined as follows: (i) asymptomatic - a person found positive for SARS-CoV-2 with no apparent signs or symptoms of disease, (ii) paucisymptomatic - a person found positive for SARS-CoV-2 with general mild symptoms (e.g. with general malaise, low grade fever, tiredness etc.) but no clear signs of disease, (iii) mild - a person found positive for SARS-CoV-2 with clear signs and symptoms of disease (e.g., respiratory disease) but not severe enough to require hospitalisation, (iv) severe - a person found positive for SARS-CoV-2 with clear signs and symptoms of disease (e.g. respiratory disease) and severe enough to require hospitalisation, and (v) critical - a person found positive for SARS-CoV-2 with clear signs and symptoms of disease (e.g., respiratory disease) and severe enough to require admission to an Intensive Care Unit (ICU).

The surveillance system captures whether the reported subject is a Health Care Worker (HCW). We have defined a HCW broadly as a person who has ever worked in the Health Care sector regardless of role, profession or current working status. The system also records whether the affected person has one of the following comorbidities: cardiovascular diseases, respiratory diseases, diabetes, immune-deficiencies, metabolic diseases, oncologic diseases, obesity, kidney diseases or other chronic diseases.

We have defined COVID-19 associated deaths as any person who has died and was confirmed to be infected with SARS-CoV-2.

Data is harmonized in a single dataset, cleaned and analysed daily to produce an infographic with main surveillance outputs. A more detailed bulletin is published bi-weekly. These outputs are publicly available on the web portal of Epidemiology of the ISS (18).

### Statistical analysis

In total, 62,843 cases were included in the analysis (data extracted on March 24, 2020). The data was summarized by age group and sex. Epidemic curves were made by date of diagnosis and of symptom onset. Cases were aggregated by region/AP of diagnosis and by province of residence for cases residing in the same region/PA of diagnosis. Attack rates per 100,000 population by region/PA were calculated using population estimates for 2019, available from the Italian National Institute of Statistics (Istituto Nazionale di Statistica; ISTAT) and adjusted using the age distribution of the Italian population as a reference. We classified the attack rates in each region as high, intermediate and low based on the inter quartile range (IQR) of the adjusted attack rates as follows: (i) high: attack rates higher than the upper limit of the IQR; (ii) intermediate – within the IQR; (iii) low – lower than the lower limit of the IQR.

Case fatality rates (CFR), not accounting for delays, were calculated by age and sex and smoothed with the locally weighted regression method. CFRs by age were also calculated by calendar period of diagnosis (i.e., before 4^th^, 4-10, 11-17, 18-24 of March). A multilevel (clustered by Region/AP) multivariable logistic model was applied to evaluate characteristics associated with death, including age group (i.e., ≤40, 40-49, 50-59, 60-69, 70-79, 80-89, 90+ years), sex, HCW status, and week of diagnosis. Adjusted odds ratios were estimated. The analyses were performed using STATA (version 16) and R (version 3.6.3). The list of the R packages used for the analysis is available in the supplementary materials.

### Transmission dynamics

As described previously (9), we define the basic reproduction number R0 as the average number of secondary cases that are generated in a fully susceptible population by a primary infector. This is an expression of the potential for transmission without any containment measure. However, once interventions are introduced or the susceptibility in the population decreases, the transmission potential at a given time t is measured as the net reproduction number *Rt*. In this paper, we estimated both *R0* and *Rt* for Italian regions in different epidemiological situations (high, intermediate and low age-adjusted attack rates), selected among those with highest data robustness. We used a previously described Bayesian approach (19–21), informed by estimates of the serial interval from contact tracing data in Lombardia (9). We defined the serial interval as the distribution of the time from symptoms onset in a primary infector and the symptoms onset in secondary cases and estimated it on average to be 6.6 days. Details are reported in the supplementary materials.

## RESULTS

As of the 24^th^ of March 2020, 62,843 confirmed cases of COVID-19 had been reported including 5,541 related deaths, and the number of reported cases was increasing (Figure 1). Locally acquired cases diagnosed at the end of February reported onset of symptoms from the 28^th^ of January onwards, indicating undetected local transmission for at least three weeks before detection. The average delay between symptom onset and diagnosis in the first month of the outbreak was 5.3 days.

**Figure 1.**
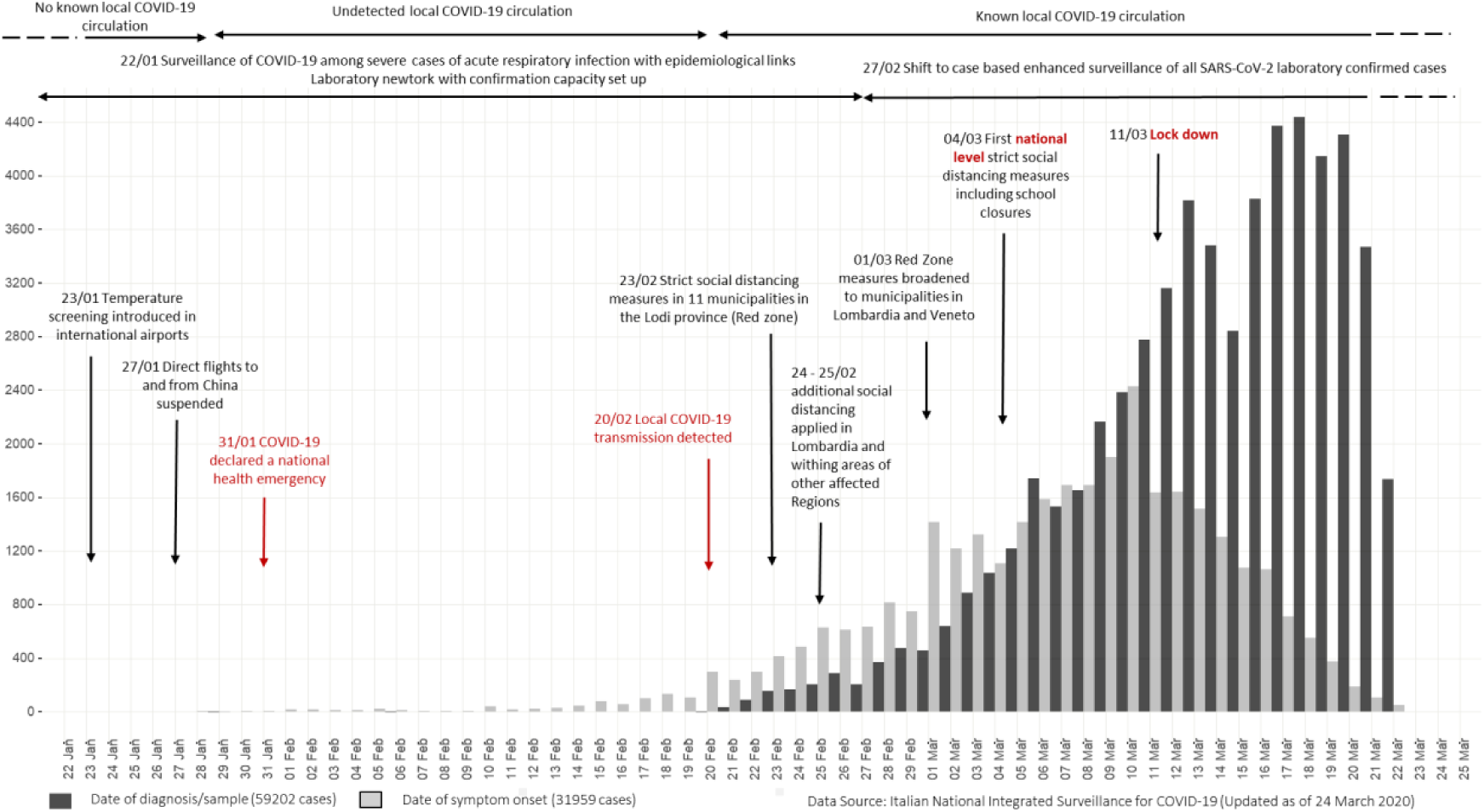
Epidemiological curves of COVID-19 cases by date of onset (blue) and date of diagnosis (green), Italy 22 January – 24 March 2020.

By March 24, 2020, all of Italy’s 21 regions and APs had reported at least one locally acquired case of COVID-19. The country had high incidence areas with sustained local transmission (mainly in the north), low incidence areas with limited but growing numbers of locally acquired cases of infection and regions with intermediate incidence (Figure 2). Overall, 98% of 29,938 COVID-19 cases diagnosed in Lombardia, were among people residing in this Region. Among the remaining 2% for which the residence of the case was known, most (94 cases) resided in the neighbouring region of Emilia Romagna. The index case of the outbreak was not found, and no clear chains of transmission were identified linking initial cases in newly affected regions/AP.

**Figure 2.**
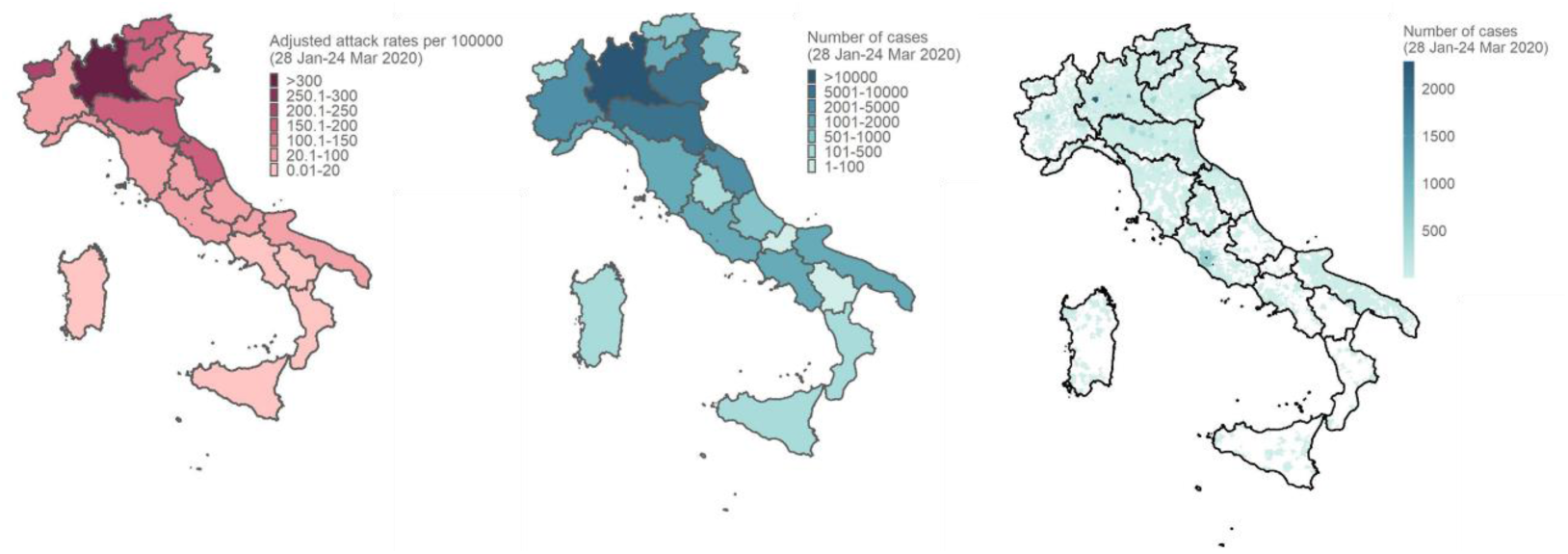
COVID-19 attack rates per 100,000 population (age-adjusted) by region/AP of diagnoses (a), number of cases, by region/AP of diagnosis -data available for 62,843 cases (b) and number of cases by municipality of residence when in the same region of diagnoses-data available for 59,128 cases (c), Italy, 24 March 2020.

Age –adjusted attack rates per 100,000 were classified as high in Lombardia (crude attack rate 303.6 and age-adjusted attack rate 305.2 per 100,000), Valle d’Aosta, Trento, the region of Emilia-Romagna bordering Lombardia and in the Marche region in central Italy. Age –adjusted attack rates were classified as intermediate in Bolzano/Bozen, Veneto, Piemonte, Liguria, Friuli-Venezia Giulia, Abruzzo, Toscana, Umbria, Molise and Puglia. In Lazio, Campania, Sardegna, Calabria, Sicilia and Basilicata age –adjusted attack rates were classified as low (Figure 2 and Table 1, supplementary material).

**Table 1.** Estimated epidemic doubling time and R0 in selected Italian Regions (data up to March 24, 2020).

| Region | Adjusted AR Classification | Doubling time (days) | $R_0$ |
| --- | --- | --- | --- |
| **Lombardia** | High | 2.7 (95%CI: 2.2-3.5) | 2.96 (95%CI: 2.73-3.17) |
| **Veneto** | Intermediate | 3.2 (95%CI: 2.5-4.2) | 2.51 (95%CI: 2.18-2.86) |
| **Emilia-Romagna** | High | 2.7 (95%CI: 2.3-3.3) | 2.84 (95%CI: 2.57-3.13) |
| **Toscana** | Intermediate | 3.2 (95%CI: 2.3-5.2) | 2.50 (95%CI: 2.18-2.83) |
| **Lazio** | Low | 2.9 (95%CI: 2.2-4.3) | 3.00 (95%CI: 2.68-3.33) |
| **Puglia** | Intermediate | 2.9 (95%CI: 2.2-4.3) | 2.61 (95%CI: 2.13-3.13) |

Most affected cases were male (58%) and the median age of cases was 63 years. Notably, 5,760 cases were reported among health care workers (median age 49 years, 35.5% male).

Among cases with known age, clinical severity was reported for 15,940 cases. As shown in Figure 3, the proportion of severe cases in under 7 years of age was below 20%. This proportion decreased to 1.5% in the 7-19 year age group, then increased again gradually to 31.9% in the age group 80-89 years. Critical severity was reported in cases aged 20 years and above, reaching 8% in the 60-69 year age segment (Table 2, supplementary material).

**Table 2.** Reported clinical severity of COVID-19 cases (n= 15,940) by Age Group, Italy, 24 March 2020.

| Age group | Asymptomatic |  | Paucisymptomatic |  | Mild |  | Severe |  | Critical |  | Total |
| --- | --- | --- | --- | --- | --- | --- | --- | --- | --- | --- | --- |
|  | N | % | N | % | N | % | N | % | N | % |  |
| 0-1 | 5 | 7.9 | 12 | 19.0 | 39 | 61.9 | 7 | 11.1 | 0 | 0 | 63 |
| 2-6 | 8 | 25.8 | 7 | 22.6 | 11 | 35.5 | 5 | 16.1 | 0 | 0 | 31 |
| 7-19 | 39 | 28.5 | 39 | 28.5 | 57 | 41.6 | 2 | 1.5 | 0 | 0 | 137 |
| 20-29 | 107 | 18.9 | 136 | 24.1 | 263 | 46.5 | 53 | 9.4 | 6 | 1.1 | 565 |
| 30-39 | 127 | 12.4 | 265 | 25.8 | 484 | 47.1 | 135 | 13.1 | 17 | 1.7 | 1,028 |
| 40-49 | 201 | 10.3 | 382 | 19.7 | 939 | 48.3 | 352 | 18.1 | 70 | 3.6 | 1,944 |
| 50-59 | 278 | 8.6 | 486 | 15.1 | 1523 | 47.3 | 771 | 23.9 | 163 | 5.1 | 3,221 |
| 60-69 | 161 | 5.0 | 310 | 9.7 | 1560 | 48.6 | 919 | 28.6 | 258 | 8.0 | 3,208 |
| 70-79 | 99 | 3.0 | 250 | 7.7 | 1696 | 52.2 | 954 | 29.4 | 247 | 7.6 | 3,246 |
| 80-89 | 68 | 3.3 | 181 | 8.7 | 1078 | 52.0 | 661 | 31.9 | 84 | 4.1 | 2,072 |
| >=90 | 30 | 7.1 | 79 | 18.6 | 196 | 46.1 | 113 | 26.6 | 7 | 1.6 | 425 |
| **Total** | **1,123** |  | **2,147** |  | **7,846** |  | **3,972** |  | **852** |  | **15,940** |

**Figure 3.**
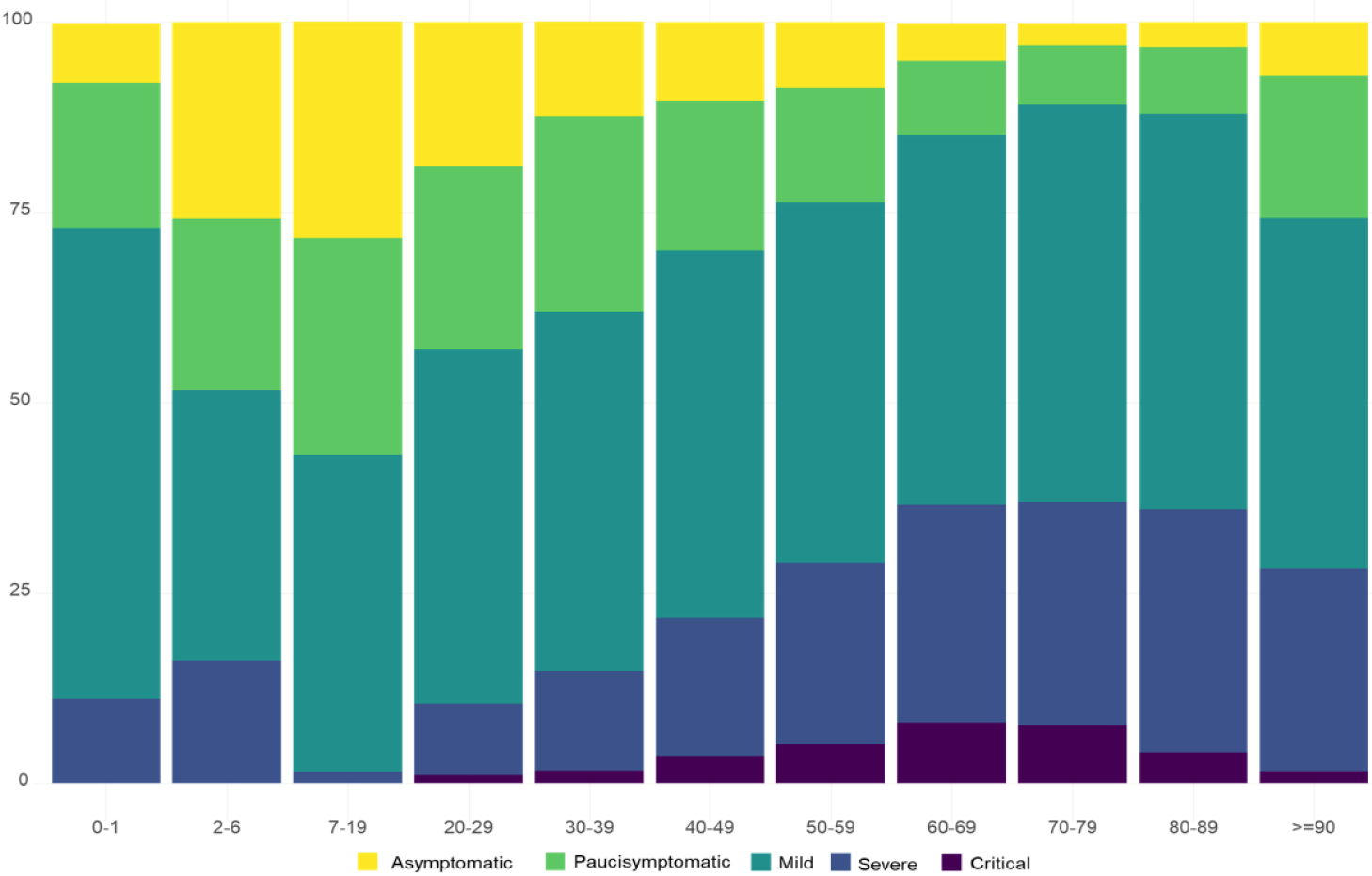
Reported clinical severity of confirmed COVID-19 cases, Italy (N=15 940)

Among all cases, 36,893 (58.7%) were reported to have been managed at home and 9,719 (15.5%) were hospitalised. For 16,231 (25.8%) cases this information was not available. As shown in Figure 4 (a), the proportion of hospitalised COVID-19 cases decreased from 0-1 to 7-19 years of age and increased progressively from 20-29 to 70-79 years of age when it appeared to stabilize. Figure 4 (b) shows, among all reported hospitalised COVID-19 cases with known unit of admission and age (N= 7,931), the proportion of patients admitted in ICU vs those admitted to any other hospital unit. Overall, the ICU admission rate was 11.2% considering only cases for which admission hospital unit was reported. Among all cases reported to have been hospitalised, the ICU admission rate was 9.1%. ICU admissions were reported from the 20-29 age group onwards, increasing in proportion until ages 60-69 years. The proportion decreases in older age groups.

**Figure 4.**
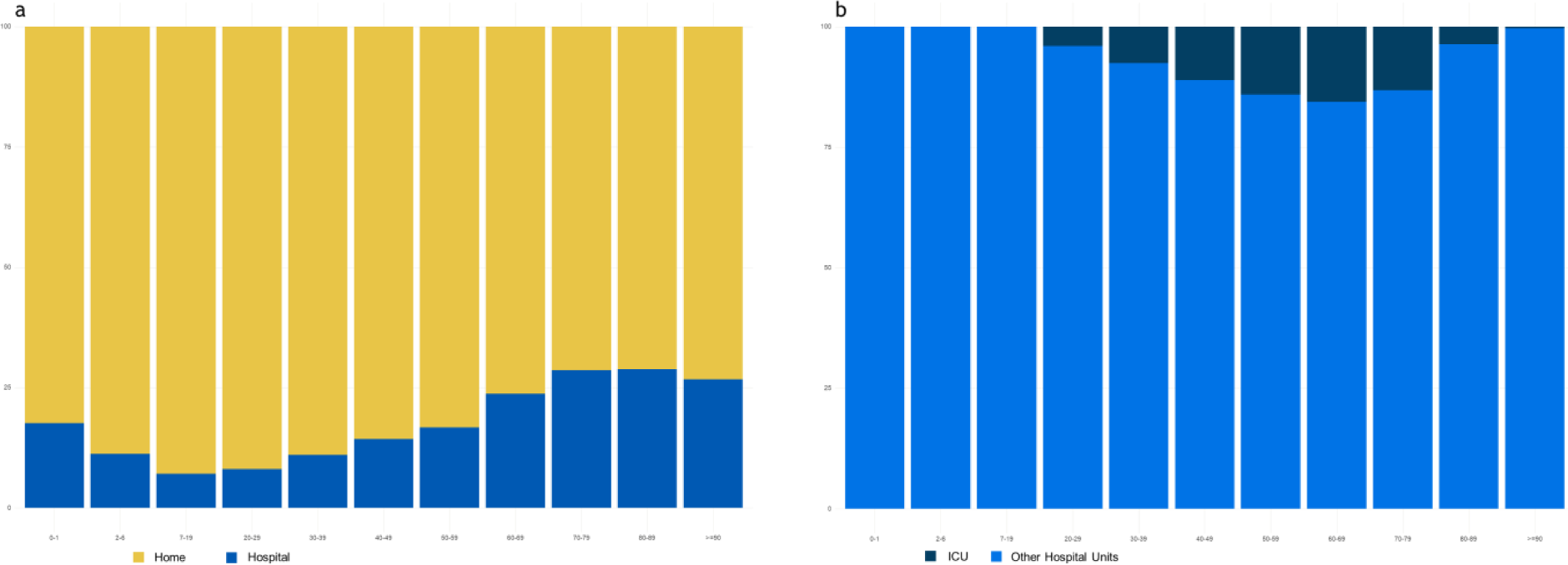
Reported proportion of COVID-19 cases by home vs hospital management (N= 46,506) (a) and proportion of hospitalized COVID-19 cases (N= 7,931) by ICU vs other Unit of hospitalisation, by age group (b), Italy, 24 March 2020.

Of the 5,541 reported COVID-19 associated deaths, 49% occurred in cases aged 80 years or above with an overall crude case fatality rate (CFR) of 8.8% (Table 3, supplementary material). Overall, 15 deaths were reported in HCW.

**Table 3.**
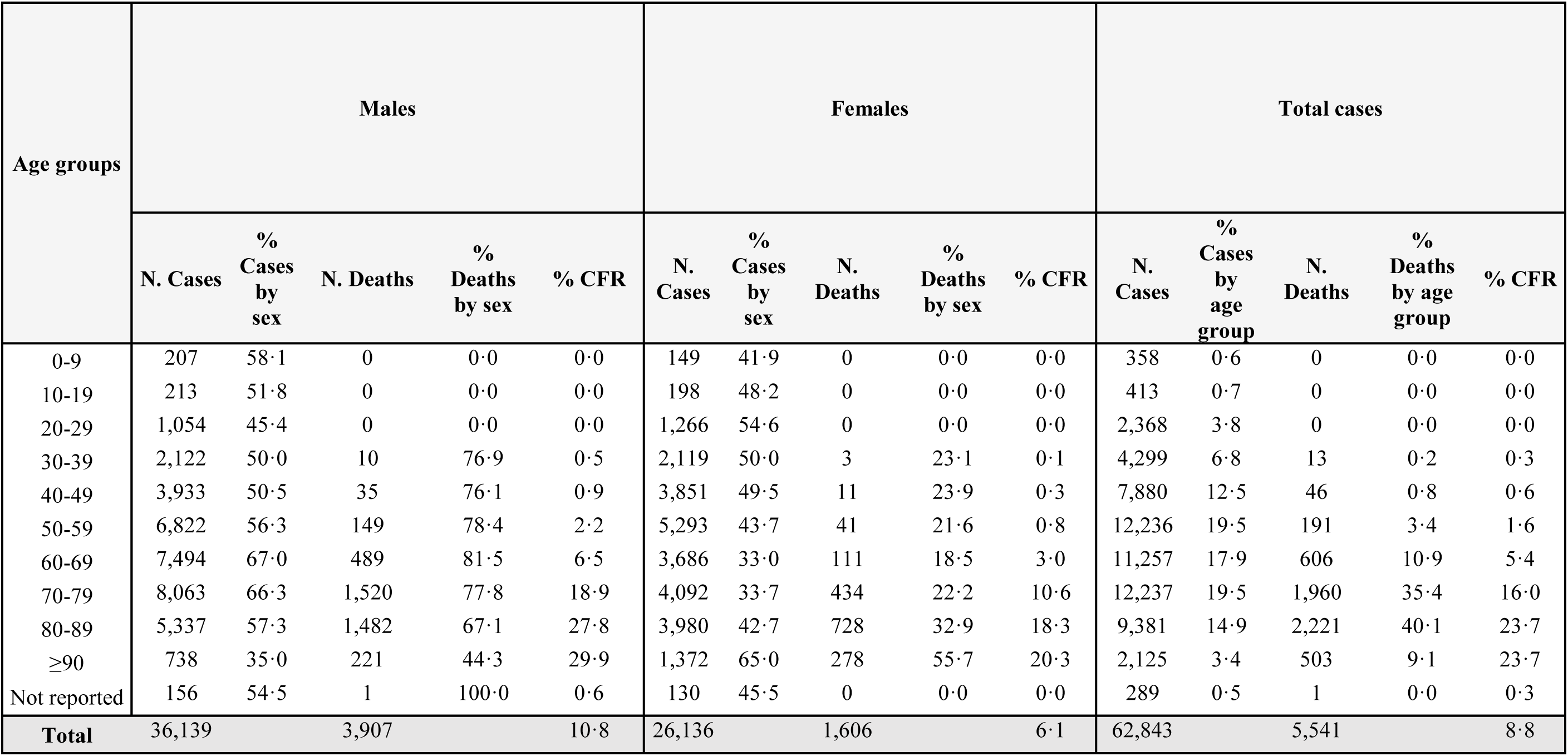
Distribution of diagnosed COVID-19 cases (n= 62,843) and related deaths (n= 5,541) by age and sex.

Until March 24, 2020, no deaths were reported among cases aged less than 30 years. Overall, 68% of the people who died were reported to have at least one co-morbidity. Figure 5 shows the CFR reported as of the 24^th^ of March 2020 by single age of diagnosed cases stratified by sex smoothed using locally weighted regression curves. There was a strong association of the CFR with age. Women had a lower CFR at each age point.

**Figure 5.**
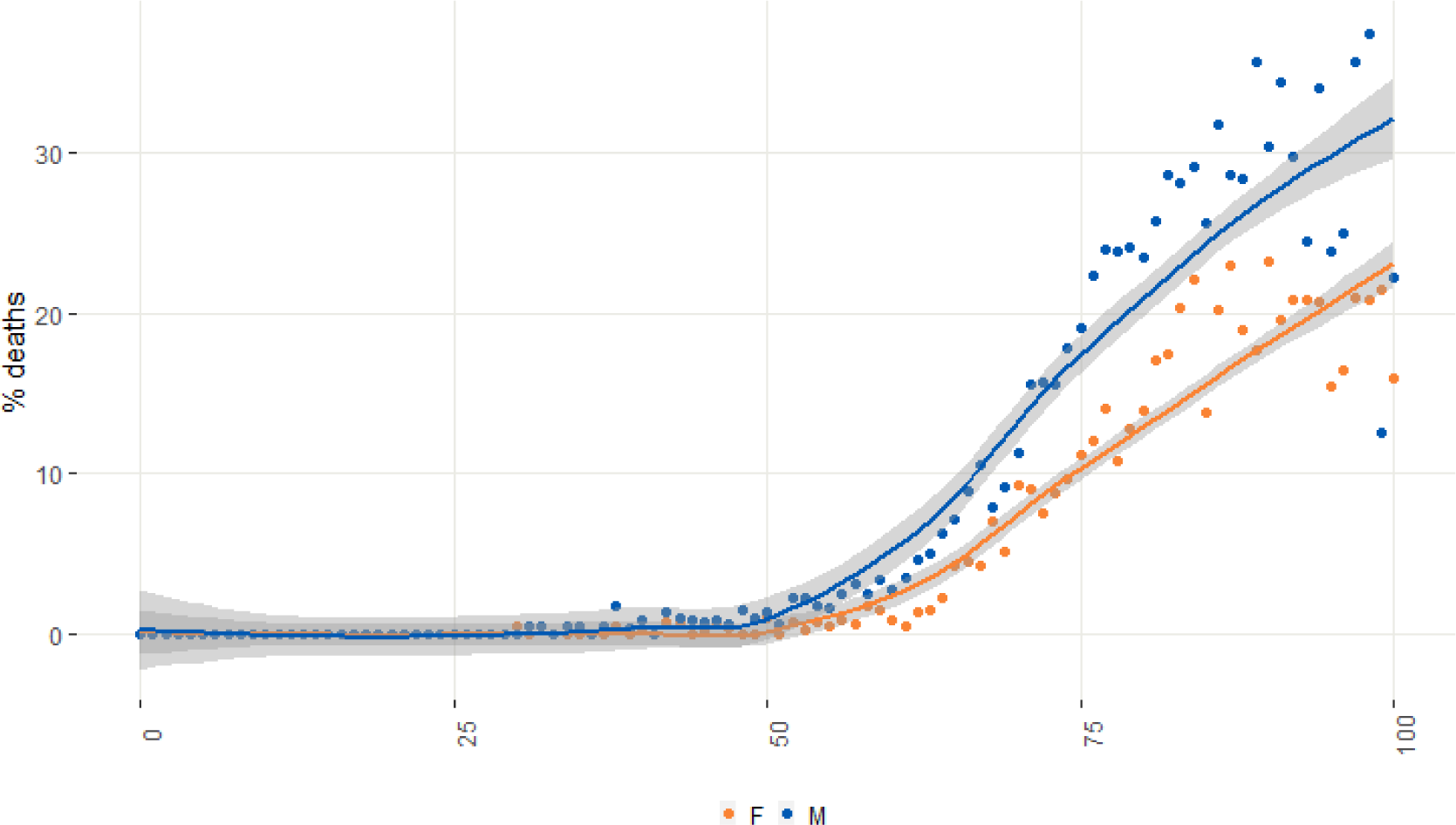
Case fatality rate by age at diagnosis and sex (dots (blue for males, reds for females) represent rates by single age, locally weighted regression curves (band width=0.8) show the estimated trend.

When stratifying by age group and calendar period of diagnosis we confirmed the age effect on the CFR but also showed that CFR increases with time since diagnosis, with an overall CFR of 19% for people diagnosed with COVID-9 before the 4^th^ of March (Table 4, supplementary material).

**Table 4.**
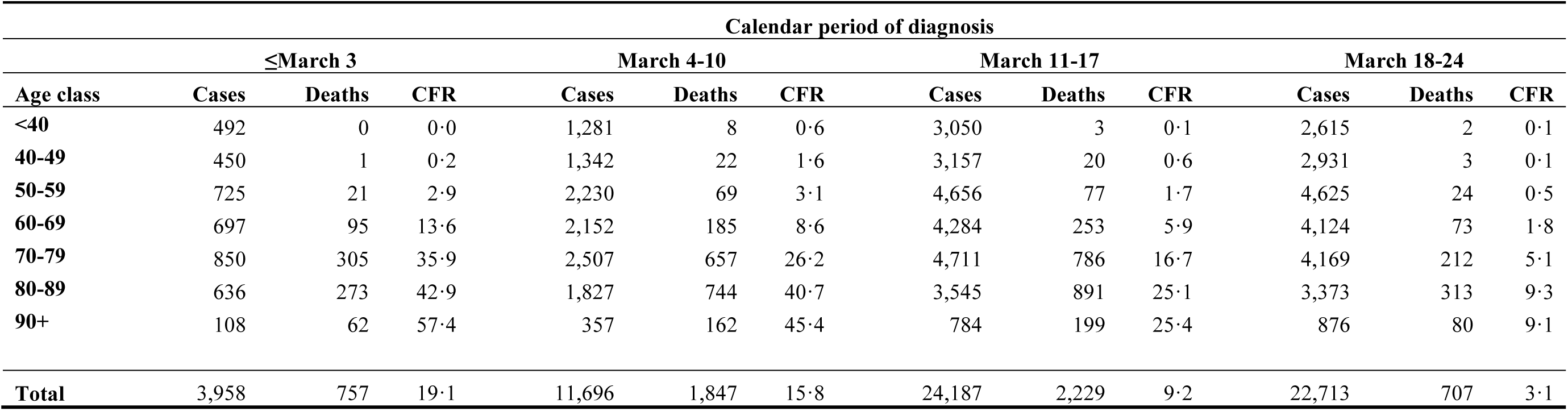
Case fatality rate by age group and calendar period of COVID-19 diagnosis.

After adjusting for age, sex, HCW profession, and calendar period of diagnosis, we estimated a higher adjusted odds ratios (AOR) of death with increasing age decade and a higher AOR for males compared to females. Health care workers (HCW) diagnosed with COVID-19 had lower AOR of death compared to the non HCW (Table 5, supplementary material). We performed additional models evaluating a possible interaction between sex and age group and between being a HCW and both age and sex. However, these did not provide a better fit to the data (p=0.06, p=0.91, p=0.26 log-likelihood ratio test, respectively). Similar results were found when restricting the analysis to cases aged 20-70 years old.

**Table 5.** Adjusted odds ratios (from multilevel logistic model clustered on reporting regions/autonomous provinces) of death in Italian COVID-19 reported cases to the Italian National surveillance (extracted March 24, 2020)

|  |  | Odds Ratio | 95% CI |  | p-value |
| --- | --- | --- | --- | --- | --- |
| **Age (years)** | *<40 (ref)* | 1.00 | - |  |  |
|  | *40-49* | 3.55 | 1.92 | 6.59 | <0.001 |
|  | *50-59* | 8.52 | 4.85 | 14.99 | <0.001 |
|  | *60-69* | 25.67 | 14.78 | 44.58 | <0.001 |
|  | *70-79* | 81.92 | 47.36 | 141.71 | <0.001 |
|  | *80-89* | 154.74 | 89.44 | 267.70 | <0.001 |
|  | *90+* | 211.45 | 121.16 | 369.02 | <0.001 |
| **Sex** | *Males vs females* | 1.79 | 1.68 | 1.92 | <0.001 |
| **Health care worker** | *Yes vs no/not indicated* | 0.11 | 0.07 | 0.19 | <0.001 |
| **Calendar week of diagnosis (<9 up to 12)** | *(per 1 week increase)* | 0.54 | 0.52 | 0.56 | <0.001 |

61,979/62,843 (98.6%) cases included in this analysis

We estimated the transmission patterns for six Italian Regions (Lombardia, Veneto, Emilia-Romagna, Toscana, Lazio, and Puglia) with adjusted AR classification ranging from low to high. These six Italian Regions are characterized by highly different epidemic trajectories. The variability is also clearly visible in terms of epidemic doubling time, which varies between 2.7 days (95%CI: 2.3-3.3) in Emilia Romagna and 3.2 days (95%CI: 2.5-4.2) in Veneto (Table 1), and basic reproductive numbers, which lie in the range 2.13-3.33.

In Lombardia, we estimated the net reproduction number (Rt) to be above the epidemic threshold since late January 2020 (Figure 6). In February, the Rt started to oscillate reaching maximum values around 3 over the week from February 17 to 23. Starting from February 24, with the enforcement of a quarantined area around the most affected municipalities of the region, Rt was estimated to follow a constantly decreasing trend. The second and third most affected regions in February (Veneto and Emilia Romagna) show an increasing trend of Rt until about February 24 (Figure 6). At that time a few tens of cases had been detected in those regions.

**Figure 6.**
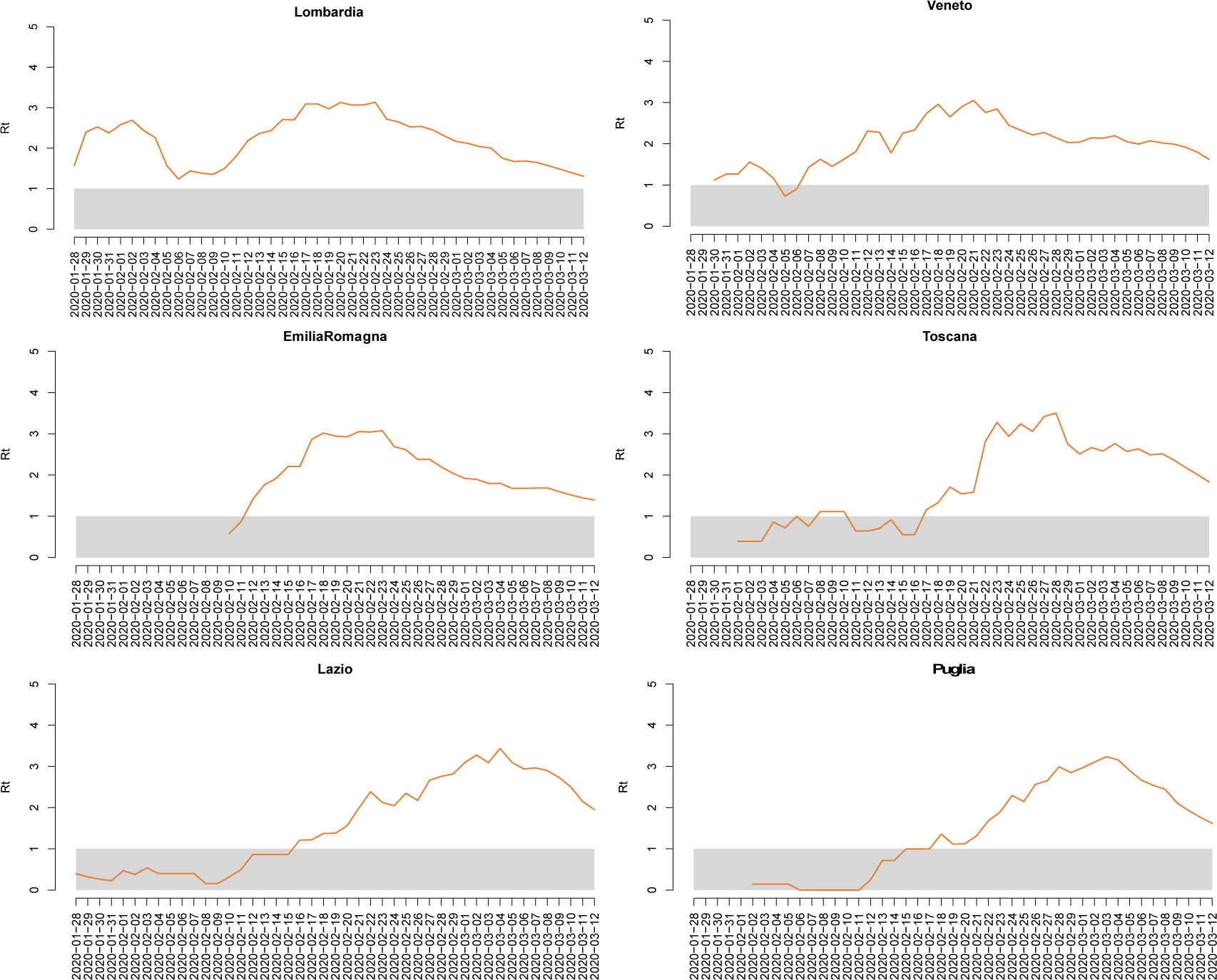
COVID-19 estimated Rt and 95%CI in selected Italian regions, February-March 2020, over a 4-day moving average.

The transmission patterns of Toscana, Lazio, and Puglia are markedly different. In Central and South Italy, where Tuscany, Lazio, and Apulia are located, the epidemic spread was largely undetected until early March. After an initial increase, Rt remained nearly constant at values around 2.5-3 until March 4-8, when blanket physical distancing measures began being implemented at national level (Figure 6).

## DISCUSSION

COVID-19 emerged in Italy with a similar clustering onset to the one described in Wuhan, China (2) with three major clusters around the cities of Codogno, Bergamo and Cremona in the Lombardia region in northern Italy (9). Subsequently, cases spread across the country with more sustained transmission in neighbouring regions in the north and in the central region of Marche. We estimated R0 in the range of 2.13-3.33 in different Italian regions with doubling time estimated between 2.7 and 3.2 days. This, alongside the short serial interval of COVID-19 in Italy (6.6 days on average) and the fact that the outbreak was detected amid ongoing transmission that had started at least three weeks earlier, explains the rapid case increase and geographical spread. It also explains the inability to identify the index case and clearly trace the initial spread of infection across the country.

Notably, in northern regions (Lombardia, Veneto and Emilia-Romagna), the net reproduction number Rt shows a marked decreasing trend since the identification of the first cases in late February. The observed decrease of Rt in northern regions is possibly due to increased population awareness and to the early effect of interventions. In other parts of the country, such as in central (Toscana, Lazio) and southern regions (Puglia), transmission was largely undetected until the first days of March, with Rt decreasing after the initial implementation of national blanket physical distancing measures.

Cases were for the most part detected in the same region/AP in which they reside. Most initial cases that did not reside in Lombardia resided in neighbouring Emilia-Romagna. This could be related to the fact that initial clusters occurred in industrial hubs with strong cross-border connections with neighbouring provinces in other regions, possibly favouring the rapid geographical spread of the infection in the north of the country.

Reported clinical severity was confirmed to be lowest among children, and in particular in the 7-19 years age group. The proportion of severe and critical cases increases with age until age 80 or above. The slight decrease in the proportion of critical and severe cases, and of deaths, in the higher age groups might be due to the demographic structure of the population with a higher female to male ratio among older people (22).

The proportion of hospitalized COVID-19 cases appears to follow a similar trend with age. Hospitalisation rates decreased from infancy to childhood and adolescence to increase again progressively with age among adults stabilizing from the 70-79 years age group.

Overall, the ICU admission rate based on reported data ranged between 9.1% and 11.2% depending on the denominator used. Both rates are much higher than the reported 4% in 16 European Union countries disclosed by the European Centre for Disease Prevention and Control (ECDC) (23). We are unable at this stage to speculate whether the reasons behind this difference are related with hospitalisation policies and practices or whether there are other factors at play. However, this observation confirms recent studies in Lombardia that highlight the potentially catastrophic effects of an uncontrolled COVID-19 epidemic on the healthcare system (8,9). Consistently with reported clinical severity, ICU admissions were notified starting from the 20-29 years age segment, suggesting the potential for critical disease also among young adults. The proportion of COVID-19 related ICU admissions, increases until the 60-69 years age segment to decrease in older ages apparently more rapidly than the reported stabilization of more severe disease in the same age groups. This pattern, however, might simply reflect lack of completeness of the surveillance data available at this stage. Further analysis on a larger number of records is required to confirm this finding.

As shown previously in China (24), we also demonstrated worse outcomes in older males with comorbidities. Male sex and older age are independent risk factors for COVID-19 related deaths. The CFR of 8.8% documented overall in Italy, is higher than that observed in other countries. As recently described, this could be in part explained by the demographic structure of the Italian population (25). However, other aspects such as testing policies and the surveillance system characteristics, including the choice in Italy to count only laboratory confirmed cases and to define associated deaths in a very inclusive manner, may also play a role in making initial case fatality data poorly comparable across countries. It is however noteworthy to stress that the early phase of the COVID-19 epidemic in Italy is characterized by a large number of cases with a short follow-up time since diagnosis. This implies that the overall CFR is currently largely underestimated. This was confirmed when performing an analysis by period of diagnosis. We found that those diagnosed before March 8th, 2020 had an overall CFR of around 20%. Future studies with longer follow-up will better clarify this aspect including studies evaluating overall population excess mortality, which will also be more comparable across countries.

Surveillance data shows clearly a very high number of COVID-19 cases among HCW, underlining the fact that SARS-CoV-2 can be easily spread in health care contexts and the importance of strong infection prevention and control practices. In Italy, as described in other countries (26), nursing homes and long term care facilities have emerged as particularly fragile environments in which infection can spread very rapidly with potentially more severe outcomes due to the vulnerability of the hosted populations (27). Affected HCW, compared with the affected general population, are on average younger and more frequently female. Considering the predominance of female professionals in working age in the Italian health sector, this is the distribution that would be expected in professionally exposed groups. Age and sex only explains in part the lower CFR, as a lower AOR of death was observed among HCW also after adjusting for those variables and might be related to earlier detection and management.

The data collected from the Italian integrated COVID-19 surveillance system during the initial phase of the emergency presents a number of limitations mainly due to completeness challenges. For this reason, some stratifications and analysis were not shown. The lack of completeness on the presence and type of comorbidities, did not allow us to include this in the multivariable analysis of deaths in order to assess, and/or adjust for, this factor. Data on hospitalisation and ICU admissions as well as CFRs are not adjusted for the expected time for disease evolution and might therefore be under-estimated in the more recent period. Finally, the estimation of R0, Rt and the doubling time were performed in regions selected on the basis of the robustness of data considering epidemiologically diverse settings.

Even in the presence of the mentioned limitations, our analysis suggests that the SARS-CoV-2 transmission potential in Italy is decreasing, albeit with large diversities across the country. Further, we observe that as of March 8 2020, the Rt it is still above the epidemic threshold. The progressively harsh physical distancing measures enacted since then may have enhanced the decreasing trend in transmissibility as happened in China [19,20]. The surveillance system will be key to monitor the effect of the implemented policies and guide the public health response as Italy will enter the second phase of its outbreak.

## PUTTING RESEARCH INTO CONTEXT

Evidence before the widespread sustained local transmission of COVID-19 in Italy was largely based on the experience shared by China (2,3) and specific other countries in Asia, while evidence in Europe was initially limited to small local clusters in Germany, France (5,6) and the UK (7). While some commentaries have been issued regarding the evolving situation in Italy (25) and initial studies are available on the epidemic in the first affected region Lombardia (8,9) or based on different modelling approaches, there has been yet no communication to the scientific community of the Italian epidemiological situation at national level based on surveillance data. In this paper, we summarize key epidemiological findings from data on the first 62,843 confirmed COVID-19 cases in Italy, including 5,541 associated deaths, and initial findings on SARS-CoV-2 transmissibility across different regions. The added value of this study is of providing an in depth review of the first month of the Italian outbreak through descriptive and analytic epidemiology and an estimation of the R0 and Rt taking into account the diversity of transmission across the country. We believe that the evidence collected in Italy, with a demographic structure and health system organization that differs from those of other countries that had reported sustained disease transmission, can be of support to countries currently experiencing initial or escalating COVID-19 transmission.

## Data Availability

All data referred to in the manuscript are collected through the national integrated surveillance system for COVID-19 in Italy

## Supplementary materials

**Figure 1.**
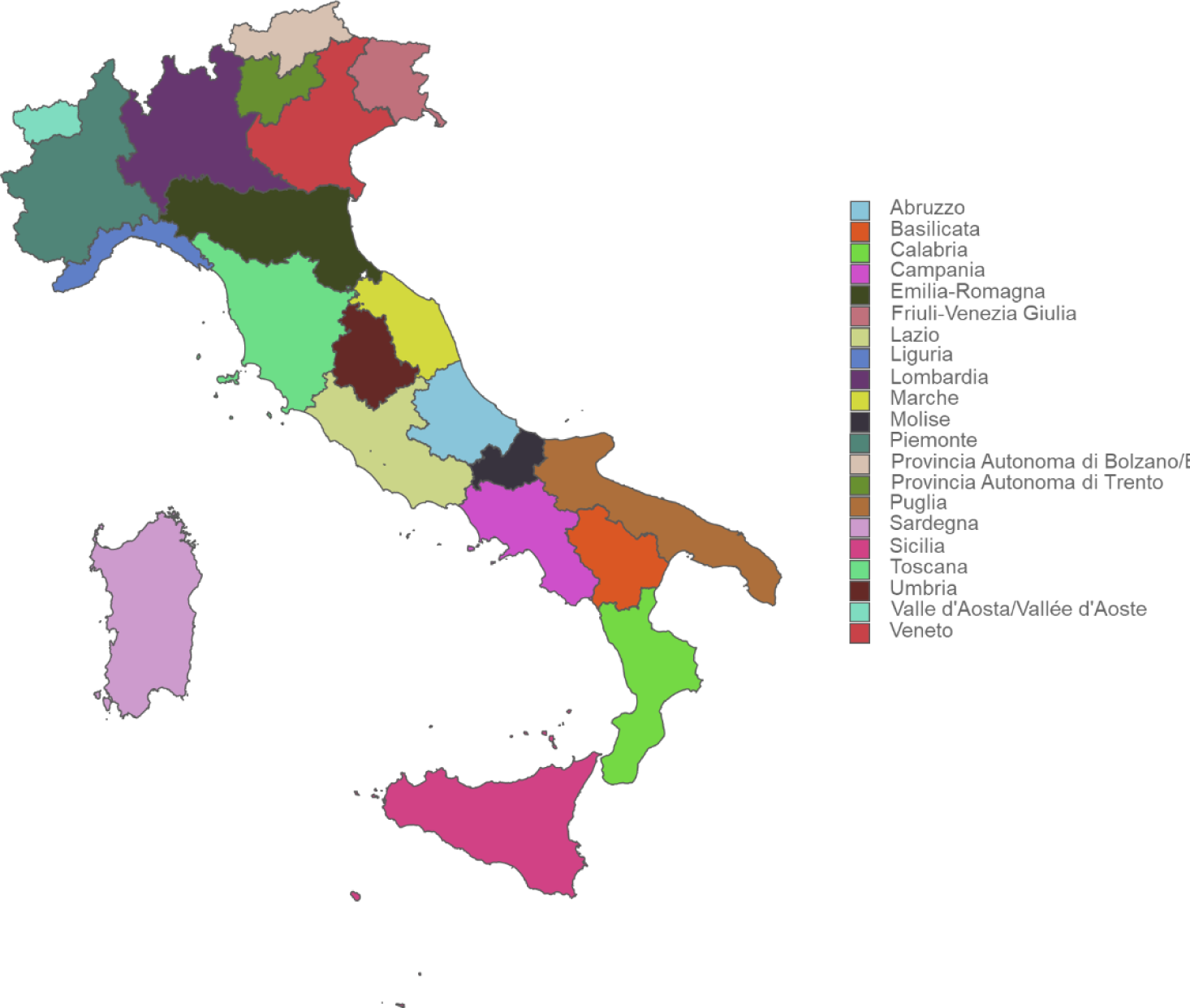
Map of the 21 Italian regions and autonomous provinces.

**Table 1.** Distribution of diagnosed COVID-19 cases (n= 62,843), crude and adjusted Attack Rates, by Region/AP, Italy, 24 March 2020.

| Region/AP | Total number of cases | Crude attack rates (per 100,000) | Adjusted attack rates (per 100,000) | Adjusted attack rate classification |
| --- | --- | --- | --- | --- |
| **Lombardia** | 30,566 | 303.6 | 305.2 | **high** |
| **Valle d'Aosta/Vallée d'Aoste** | 307 | 227.6 | 224.8 | **high** |
| **Provincia Autonoma di Trento** | 1,002 | 185.2 | 189.1 | **high** |
| **Emilia-Romagna** | 8,489 | 190.3 | 185.7 | **high** |
| **Marche** | 2,699 | 175.7 | 167.7 | **high** |
| **Provincia Autonoma di Bolzano/Bozen** | 781 | 146.3 | 155.5 | **intermediate** |
| **Veneto** | 5,948 | 120.7 | 120.2 | **intermediate** |
| **Piemonte** | 4,201 | 96.3 | 91.7 | **intermediate** |
| **Liguria** | 1,288 | 81.3 | 73.4 | **intermediate** |
| **Friuli-Venezia Giulia** | 816 | 66.7 | 63.6 | **intermediate** |
| **Abruzzo** | 676 | 51.5 | 50.5 | **intermediate** |
| **Toscana** | 1,761 | 47.0 | 45.4 | **intermediate** |
| **Umbria** | 239 | 26.8 | 26.6 | **intermediate** |
| **Molise** | 72 | 26.5 | 25.5 | **intermediate** |
| **Puglia** | 1,004 | 24.7 | 25.2 | **intermediate** |
| **Lazio** | 1,222 | 20.7 | 20.9 | **low** |
| **Campania** | 1,021 | 17.1 | 18.0 | **low** |
| **Sardegna** | 223 | 13.3 | 12.8 | **low** |
| **Calabria** | 176 | 8.8 | 9.1 | **low** |
| **Sicilia** | 342 | 5.1 | 5.2 | **low** |
| **Basilicata** | 10 | 2.8 | 2.8 | **low** |
| **Total** | **62,843** |  |  |  |

## ESTIMATION OF THE REPRODUCTION NUMBER

The basic reproduction number R0 represents the average number of secondary cases generated by a primary infector in a fully susceptible population. In general terms, when R0 is larger than 1 the infection may spread in the population and the larger R0 the larger effort required to control the epidemic. Once the number of susceptible individuals declines, the transmission potential of the disease at a given time t is measured in terms of the net reproduction number R(t). The net reproduction number is useful to track the effectiveness of performed control measures and other factors affecting the spread of the epidemic (e.g., the behavioral response of the population) over time. As soon as R(t) falls below 1, the epidemic starts to decline.

To estimate R(t), we use the same methodology presented in reference [1-3]. We assumed that the daily number of new cases (date of symptom onset) with locally acquired infection L(t) can be approximated by a Poisson distribution according to the equation

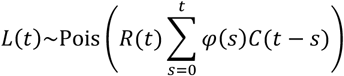

where

- C(t), with t from 1 to T, is daily number of new cases (date of symptom onset);
- R(t) is the net reproduction number at time t;
- *φ*(*s*) is the distribution of the generation time (corresponding to the distribution of the serial interval) calculated at time s. From the analysis of 90 observations of individual serial intervals in 55 clusters, the distribution of the serial interval was estimated to follow a gamma distribution with mean 6.6 days (percentiles 2.5^th^ and 97.5^th^ of the distribution: 0.7-19.0) [4].

The likelihood ℒ of the observed time series of cases from day 1 to day T conditional on C(0) is thus given by

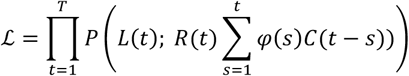

where P(k; λ) is the probability mass function of a Poisson distribution (i.e., the probability of observing k events if these events occur with rate λ).

We then used MCMC Metropolis-Hastings sampling to estimate the posterior distribution of R(t). To estimate R0, we assumed that during the period where the epidemic showed exponential growth R(t)=R0 and used the above described procedure.

